# Association of diagnosis delay and disease activity with burden on Daily life in 4150 European patients with Systemic Lupus Erythematosus

**DOI:** 10.1101/2022.11.09.22282123

**Authors:** Alain Cornet, Jeanette Andersen, Francesca Marchiori, Blanca Rubio, Philippe Mertz, Laurent Arnaud, the patient association Lupus Europe

**Author notes:** **Corresponding Author:** Pr. Laurent ARNAUD, Service de rhumatologie, Centre National de Référence des Maladies Autoimmunes et Systémiques Rares, Hôpital de Hautepierre, 1 Avenue Molière BP 83049, 67098 Strasbourg Cedex, France. |.

## Abstract

**Objective:** Despite significant improvements in diagnosis delay and treatment strategies, the burden of Systemic Lupus Erythematosus (SLE) remains high. The objective of the study was to assess the association between diagnosis delay, disease activity and burden on daily life (BoDL) in a large sample of European patients with SLE.

**Methods:** In May 2020, Lupus Europe, the European umbrella patient association for SLE, conducted a multilingual anonymous online cross-sectional study to individuals with a self-reported physician’s diagnosis of SLE living in Europe. The BoDL score was computed using 1 to 5 Likert scales on 5 domains (mobility, anxiety/depression, self-care, daily activities and pain/discomfort) and the sum was rescaled on a 0 (minimum Burden on daily life) to 100 (maximum BoDL) scale. Comparisons between independent groups were made using the Mann-Whitney test for continuous outcomes and the Chi-2 test (or Fisher’s exact test) for quantitative data.

**Results:** Data of 4,150 SLE patients from 35 European countries were analysed. Those with a diagnosis of SLE within 2 years of first symptoms had significantly lower mean BoDL scores than those diagnosed after 5 years (33.6 versus 44.0, p<0.001). The BoDL score was better in SLE patients feeling that their lupus had been under control during the last 3 months versus the others (34.0% versus 47.6%, p<0.001).

**Conclusion:** This large international study highlights the association between diagnosis delay and self-perceived disease activity with the burden of the disease on the daily life of people living with SLE. Healthcare pathways, which may accelerate diagnosis and optimize therapeutic management, are necessary to improve patients’ outcomes in SLE.

## Introduction

Systemic Lupus Erythematosus (SLE) is an autoimmune systemic disease with an incidence of 0.3 to 5.1 per 100,000 per year and a prevalence of 6.5 to 85 per 100,000 in Europe [1]. This yields an estimated 200,000 - 250,000 prevalent SLE cases across Europe. Detailed information on the characteristics and burden of SLE at the European level are largely unknown to physicians, policy makers, and lupus patients themselves. Also, due to differences in national regulations and health insurance policies, significant heterogeneity in the diagnosis and management strategies for SLE is remaining across the member states. In 2020, Lupus Europe, the European umbrella non-profit independent organisation that brings together national lupus patient organisations from across Europe, conducted a study (LWL2020) about the impact of SLE upon individuals with the disease, from the patient perspective **[2]**. The study questionnaire contained elements relating to diagnosis delay, disease activity, mobility, anxiety/depression, self-care, daily activities and pain/discomfort, providing a unique opportunity to quantify the burden of the disease and its relationship with diagnosis delay and disease activity control from the patients’ perspective.

## Patients & Methods

### Study design

From May 9 2020, until May 31, 2020, Lupus Europe conducted an on-line study amongst patients living with SLE in Europe. The detailed questionnaire and survey methodology have been previously published [2]. The study was approved by the Ethic Committee of Strasbourg Medical School (#CE-2020-109).

### Burden of disease on daily life questions & coding

A total of 5 questions provided insights on the BoDL. The first three questions (Q18 – How do you assess your mobility, i.e. your ability to walk around?; Q19 – How do you assess your ability to perform self-care tasks like washing or dressing yourself?; and Q20 – How do you assess your ability to perform normal daily activities, like studying, working, housework, leisure or participation to family life?), were coded from 1 (“fully unable” to perform) to 5 to “no problem at all”. For the other two questions (Q22 – How do you assess your level of discomfort or pain? And Q23 – Do you feel anxious or depressed?), the scoring scale was reversed as their phrasing expressed a limit rather than an ability. These two subsets of questions appeared distinct on screen, to avoid any confusion in the scales.

### Burden on daily life (BoDL) score

From these collected data, a new variable (BoDL score) was constructed. The scores of the first three questions were reversed to bring those to the same scale as the last two questions. The (1 to 5) score of each 5 questions was then added, and then converted to a 0 to 100% scale for each participant. The resulting BoDL score reflects the burden on the daily life of any given patient, ranging from 0 when no difficulties at all were reported to 100% when extreme difficulties were highlighted in all 5 domains.

### Statistical analysis

Data are presented as mean and standard deviation or counts and percentages. Comparisons between independent groups were made using the Mann-Whitney test for continuous outcomes and the Chi-2 test (or Fisher’s exact test when appropriate) for quantitative data. Two-sided p-values <0.05 were considered statistically significant. Statistical analyses were performed using STATA version 17.

## Results

### Patients characteristics

From a total of 5,922 answers (figure 1), 1547 patients were excluded because the country was out of the study scope (n=137), they did not state their country (n=271), of identified duplicate records (n=207), no lupus (n=54), drug-induced lupus (n=29), isolated Cutaneous Lupus Erythematosus (CLE) (n=342), “lupus-like disease” (*no formal lupus diagnosis*) (n=256) or because they did not mention any specific diagnosis (n=251). From the 4,375 remaining participants, 4150 (94.9%) answered all 5 questions relating to BoDL and were retained for this study. Their demographics are similar to those of the complete survey reported in LSM (table 1).

**Table 1.**
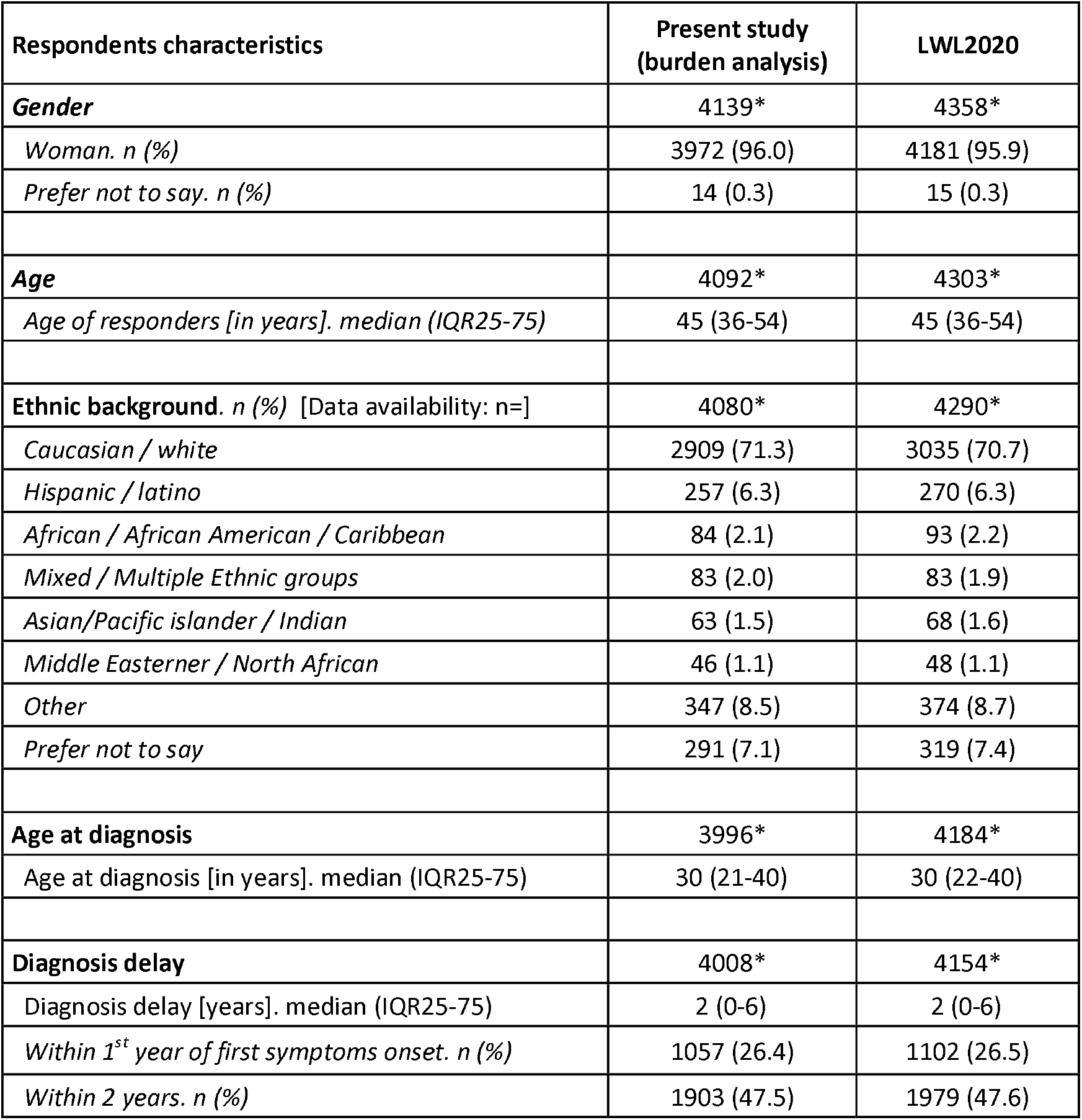
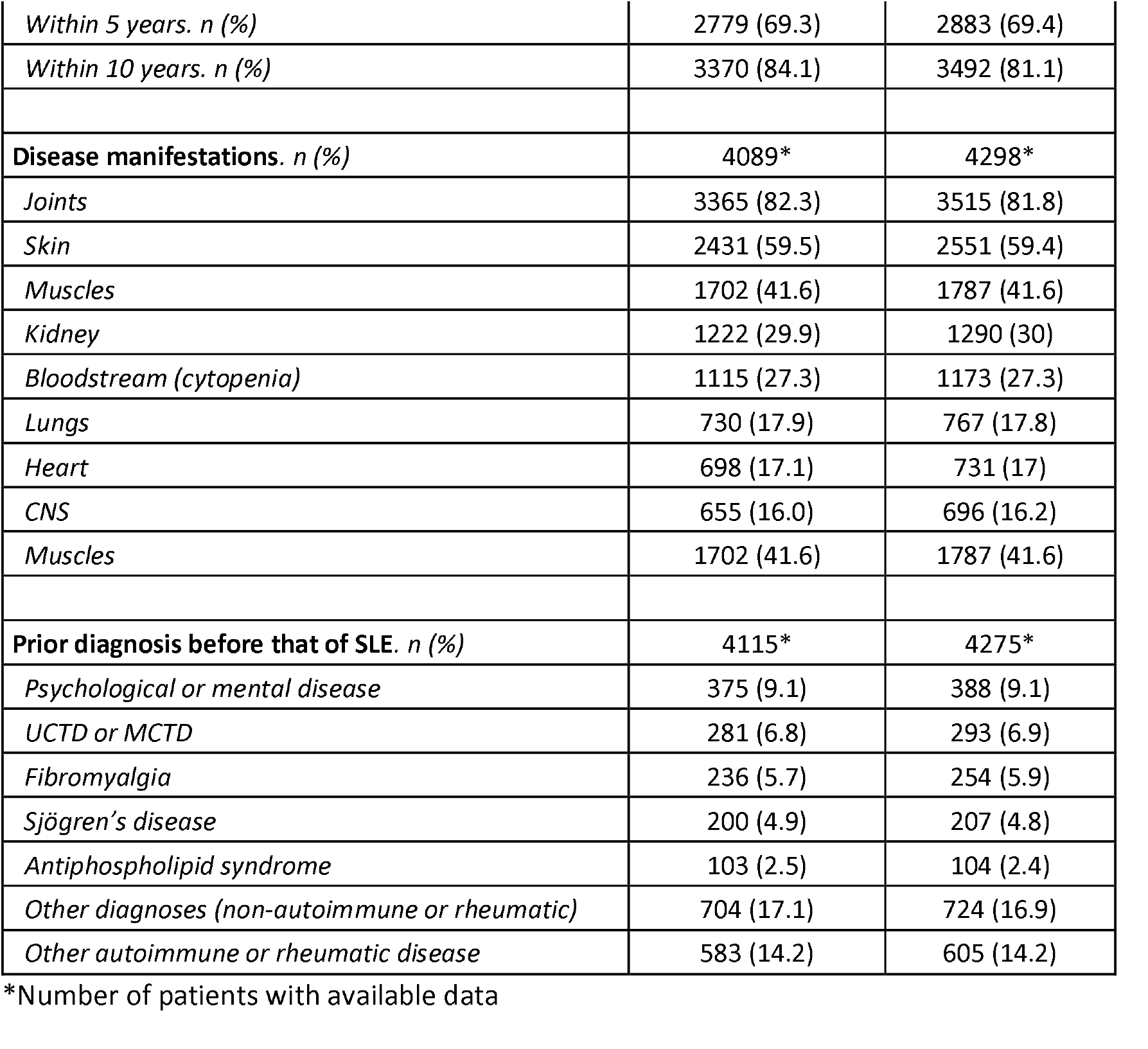
Respondent characteristics in the burden analysis study as well as in LWL2020.

| Respondents characteristics | Present study<br>(burden analysis) | LWL2020 |
| --- | --- | --- |
| **Gender** | 4139* | 4358* |
| *Woman. n (%)* | 3972 (96.0) | 4181 (95.9) |
| *Prefer not to say. n (%)* | 14 (0.3) | 15 (0.3) |
| **Age** | 4092* | 4303* |
| *Age of responders [in years]. median (IQR25-75)* | 45 (36-54) | 45 (36-54) |
| **Ethnic background. n (%)** [Data availability: n=] | 4080* | 4290* |
| *Caucasian / white* | 2909 (71.3) | 3035 (70.7) |
| *Hispanic / latino* | 257 (6.3) | 270 (6.3) |
| *African / African American / Caribbean* | 84 (2.1) | 93 (2.2) |
| *Mixed / Multiple Ethnic groups* | 83 (2.0) | 83 (1.9) |
| *Asian/Pacific islander / Indian* | 63 (1.5) | 68 (1.6) |
| *Middle Easterner / North African* | 46 (1.1) | 48 (1.1) |
| *Other* | 347 (8.5) | 374 (8.7) |
| *Prefer not to say* | 291 (7.1) | 319 (7.4) |
| **Age at diagnosis** | 3996* | 4184* |
| *Age at diagnosis [in years]. median (IQR25-75)* | 30 (21-40) | 30 (22-40) |
| **Diagnosis delay** | 4008* | 4154* |
| *Diagnosis delay [years]. median (IQR25-75)* | 2 (0-6) | 2 (0-6) |
| *Within 1<sup>st</sup> year of first symptoms onset. n (%)* | 1057 (26.4) | 1102 (26.5) |
| *Within 2 years. n (%)* | 1903 (47.5) | 1979 (47.6) |
| *Within 5 years. n (%)* | 2779 (69.3) | 2883 (69.4) |
| *Within 10 years. n (%)* | 3370 (84.1) | 3492 (81.1) |
| **Disease manifestations. n (%)** | **4089\*** | **4298\*** |
| *Joints* | 3365 (82.3) | 3515 (81.8) |
| *Skin* | 2431 (59.5) | 2551 (59.4) |
| *Muscles* | 1702 (41.6) | 1787 (41.6) |
| *Kidney* | 1222 (29.9) | 1290 (30) |
| *Bloodstream (cytopenia)* | 1115 (27.3) | 1173 (27.3) |
| *Lungs* | 730 (17.9) | 767 (17.8) |
| *Heart* | 698 (17.1) | 731 (17) |
| *CNS* | 655 (16.0) | 696 (16.2) |
| *Muscles* | 1702 (41.6) | 1787 (41.6) |
| **Prior diagnosis before that of SLE. n (%)** | **4115\*** | **4275\*** |
| *Psychological or mental disease* | 375 (9.1) | 388 (9.1) |
| *UCTD or MCTD* | 281 (6.8) | 293 (6.9) |
| *Fibromyalgia* | 236 (5.7) | 254 (5.9) |
| *Sjögren's disease* | 200 (4.9) | 207 (4.8) |
| *Antiphospholipid syndrome* | 103 (2.5) | 104 (2.4) |
| *Other diagnoses (non-autoimmune or rheumatic)* | 704 (17.1) | 724 (16.9) |
| *Other autoimmune or rheumatic disease* | 583 (14.2) | 605 (14.2) |

\*Number of patients with available data

### Diagnosis delay, disease activity and Burden of Disease Score

Of 4008 patients with available data, the diagnosis delay for SLE was reported to be <2 years in 1903 participants (47.5%), between 2 and <5 years in 1056 (26.3%) and ≥5 years in 1049 (26.2%). Patient with a diagnosis of SLE within 2 years of first symptoms had significantly better mean BoDL scores than those diagnosed after 5 years (33.6 versus 44.0, p<0.001).

A total of 2980 (71.8%) patients felt that their “lupus ha[d] been under control over the last 3 months” while 1166 (28.1%) did not (4 patients did not answer this question). Patients diagnosed within 2 years of first symptoms reported more commonly adequate disease control than those with delayed diagnosis ≥ 5 years (77.3% versus 63.3%, p<0.001).

The BoDL score was better in SLE patients feeling that their lupus had been under control over the past 3 months versus the others (34.0% versus 47.6%, p<0.001). Generally similar results were observed at the country level (table 2).

**Table 2.** BoDL scores by diagnosis delay and adequate disease control, by country.

| BoDL scores by country | Diagnosis delay <2y | Diagnosis delay ≥5y | P-value | Adequate disease control | Not under control | P-value |
| --- | --- | --- | --- | --- | --- | --- |
| Belgium | 33 | 43 | .003 | 34 | 49 | <.001 |
| Bulgaria | 35 | 45 | .003 | 34 | 49 | <.001 |
| Croatia | 32 | 37 | .20 | 34 | 41 | .08 |
| Denmark | 34 | 43 | .016 | 33 | 49 | <.001 |
| Finland | 30 | 37 | .017 | 28 | 45 | <.001 |
| France | 34 | 45 | <.001 | 35 | 46 | <.001 |
| Germany | 31 | 41 | <.001 | 31 | 45 | <.001 |
| Italy | 34 | 44 | <.001 | 35 | 46 | <.001 |
| Norway | 32 | 45 | <.001 | 36 | 45 | .004 |
| Poland | 29 | 49 | <.001 | 32 | 47 | <.001 |
| Portugal | 37 | 45 | .007 | 35 | 47 | <.001 |
| Spain | 33 | 45 | <.001 | 34 | 46 | <.001 |
| UK | 39 | 49 | <.001 | 38 | 52 | <.001 |

We identified a modest downward trend of the burden on daily life based on age (table 3), whereby people living with lupus had an overall burden on daily life increasing from 33.4% to 42.1% from age less than 25 to age 65, a loss of up to 9% points over up to 40 years.

**Table 3.** Burden of disease score, by age categories.

| Age of responders | mean BoDL score (± SD) |
| --- | --- |
| Up to 25 | 33.4 (18.6) |
| 26 to 35 | 33.6 (17.8) |
| 36 to 45 | 36.6 (18.6) |
| 46 to 55 | 40.1 (18.4) |
| 56 to 65 | 42.1 (18.5) |
| above 65 | 40.6 (20.1) |

## Discussion

The living with lupus in 2020 survey included a very large number of patients with SLE from all over Europe, and collected key elements related to the impact of the disease upon daily life. While the study did not collect HRQoL according to validated scales, this large set of data nonetheless offered a unique opportunity to assess the impact of diagnosis delay and disease activity upon the daily burden of the disease.

The LWL2020 survey highlighted that despite much progress in technology such as remote consultation, establishment and dissemination of new classification criteria, the median diagnosis delay is of 2 years and was greater than 5 years in approximately one third of patients. This is obviously a key concern for people living with lupus, because on one hand the pre-diagnosis period is known to be prone to high anxiety and difficulty to live with a series of unexplained symptoms, and on the other hand because of the concern that late diagnosis means late start of treatments, potentially leading to higher disease burden on daily life. The LWL2020 data objectivates this fact: when comparing patients groups diagnosed within 2 years of first symptoms to those diagnosed after 5 years, the burden on daily life was increased by 31% in the latter group. This trend is deemed robust as it was found across almost all European countries (table 2).

Despite current obstacles, these results highlight the importance of improving current diagnosis delay for SLE as a way to improve the burden of the disease on daily life.

The LWL2020 data also underlined the association between disease activity and the daily burden of the disease. While a majority of respondents (71.8%) felt that their “lupus had been under control over the last 3 months”, those who reported the contrary had a 40% higher burden on daily life. Again, this trend was found across almost all European countries

Among the main limitations of the study is its design as an online survey. However, the large number of respondents (>4000) and the large European coverage (35 countries) ensured a satisfactory representation of the data and offered a large international unique set of data. Another limitation is that the diagnosis delay was reported by respondents and may be prone to a memorization bias. Finally, the LWL2020 survey took place during the troubled 2020 COVID year, which may have modified the burden of the disease.

## Conclusion

This large patient survey reveals both the importance of prompt SLE diagnosis as well the relationship between disease activity and disease burden upon the daily life of European lupus patients. Further improvements should focus on reducing the diagnosis delay and identifying new therapeutic strategies for those with uncontrolled disease.

## Data Availability

All data produced in the present work are contained in the manuscript

## Acknowledgements

LUPUS EUROPE is very thankful to the volunteers that helped build and translate the survey, and then disseminate it throughout Europe, as well as all the patients that have given their time and data to help us better understand the disease.

